# Health Implications of Head Portering in Female Head Porters: A Scoping Review Protocol

**DOI:** 10.1101/2025.05.23.25326489

**Authors:** Veronica Segbedzie, Solina Richter, Pammla Petrucka

## Abstract

**Objective:** This scoping review protocol outlines the steps to conduct a scoping review to assess the health experiences of female head porters and their unique health challenges and needs. The review aims to broaden our understanding of the occupational health challenges facing head porters in sub-Saharan Africa and identify any gaps in the health literature related to this population.

**Introduction:** Head portering is an informal occupation predominantly undertaken by women and girls in sub-Saharan Africa. Female head porters often lack education and skills, which makes head portering an appealing yet laborious occupation to improve income and associated social conditions and support families left behind.

**Inclusion criteria:** Studies were included in the review if they included females of all ages engaging in head-portering activities as a livelihood in sub-Saharan Africa.

**Methods and Analysis:** The six stages of the Arksey and O’Malley (2005) framework guided the design and reporting of this scoping review. The methodology adhered to the Preferred Reporting Items for Systematic Reviews and Meta-analyses (PRISMA) guidelines for scoping reviews. Relevant articles and grey literature were identified through searches of health-related databases. The extracted data will be presented using a narrative summary to complement a data table.

**Ethics and dissemination:** The ethical approval submission for this scoping review protocol was not completed as it is not required. Formal knowledge translation and dissemination activities will include publishing findings in African-based high-impact journals and conference presentations to reach relevant audiences. The final recommendations for practice, policy, and research will be shared and distributed through toolkits for healthcare professionals and stakeholder information leaflets.

**Registration:** The review has been registered with Open Science Framework Registries: https://osf.io/c3q7as

**Strengths and Limitations:** This scoping review explores the literature on the health implications of head portering among female head porters, encompassing a broad range of studies and evidence. It aims to identify research gaps, highlight key health concerns requiring further investigation, and serve as a foundation for future reviews and primary studies. However, this review does not include a formal quality assessment of the included studies, which may impact the reliability of the findings. Due to its broad scope, detailed analyses of specific health issues may be limited. Additionally, the risk of selection and interpretation bias may constrain the depth of analytic synthesis, reducing its applicability for precise policy and practice recommendations.

## Introduction

The rapid urbanisation in sub-Saharan Africa has led to increased socio-economic inequalities and a growing burden of disease. This trend has significant health consequences, heightening the vulnerabilities of marginalised groups residing in urban centres. In contemporary migration patterns within this region, women migrate to urban areas for economic opportunities to enhance their financial capacity and improve their living conditions (11–13, 36). In urban centres, migrant women participate in commerce, often within the informal sector, which employs over 70% of the labour force (17, 28; 49). Head portering is a common informal occupation predominantly undertaken by women and girls in urban areas. This work entails carrying heavy loads on their heads over long distances, frequently in challenging environmental and socio-economic conditions. The work is typically well organised and can be lucrative, enabling women to support their families, create job opportunities for low-income earners, and provide access to food resources for those unable to afford it through established credit systems in their communities (20, 35). Head portering among females presents complex intersections of socio-economic challenges, gender dynamics, and health implications. The health effects of head portering are increasingly concerning due to the physical, mental, and social challenges associated with this occupation (46).

Understanding the occupational health challenges of head portering impacting the well-being of head porters is essential for developing targeted interventions designed to improve their quality of life. This scoping review aims to map and synthesise existing literature on the health experiences of head porters in marketplaces across sub-Saharan Africa. It will identify gaps in the current literature, summarise known head porters’ health challenges, and suggest future research directions.

## Background

Despite global efforts to enact constitutional safeguards for all, women and girls remain vulnerable, often limiting their access to material and economic resources (21, 22, 48, 56). In sub-Saharan Africa, women constitute a significant portion of the informal labour force in urban areas (6). Informal work is unregulated and typically involves low wages, poor working conditions, and limited access to social protection, including health care and pension schemes (55). Migration to engage in informal work is one strategy young girls and women utilise to cope with worsening socio-economic problems brought on by historical, political, demographic, economic, and ecological pressures (10, 29, 35, 36).

Head portering has been a longstanding informal occupation in urban centres across sub-Saharan Africa, with a significant portion of female internal migrants and marginalised individuals engaging in this informal labour (11, 46). This occupational activity has been widely documented in Ghana, Nigeria, Mali, and Burkina Faso, where rural-urban migration patterns are common, particularly among young women and girls seeking income (35). Head porters may be referred to by different names depending on the country and the local language; for example, Alabaru in Nigeria and Kaya Yei in Ghana (5, 7).

Head porters play a vital role in commercial centres’ economic development by transporting heavy goods in areas where it is not possible to move goods by vehicle (5). They manually transport goods by carrying loads on their heads for customers or traders in exchange for a small fee. The income earned through this precarious work returns to the national economy through payment for daily market tolls and provides access to community facilities, including accommodation, use of bathrooms, and water (43). It also serves as a vital source of income and supports family expenses in the villages of origin, including school fees and medical bills for dependents (3, 4).

This work is physically demanding, poorly remunerated, and typically undertaken by individuals from disadvantaged or marginalised communities, often female rural migrants seeking better economic opportunities in cities. While head portering is associated with women and girls, male head porters exist in specific contexts (6). Cart pushing by male porters enables them to work in the same settings and carry heavier loads over longer distances, earning more money and reducing the risk of injury (5). Hence, gender impacts the types of work done in commercial areas by women, as it reflects social hierarchies and power relations regarding how workspaces are negotiated and used (52). In addition, these female migrants experience negative public perceptions due to their ethnicity and the tendency to engage in circular migration, which limits their ability to take full advantage of urban centres’ affluence and social welfare (3, 35). As a result, carrying loads on their heads becomes the primary option for women who rely on physical body strength whilst earning less compared to men who often use push wooden carts and wheelbarrows to transport loads (1, 5). Despite their critical role in urban economics, the occupational hazards impacting the health of female head porters remain understudied, highlighting the need for targeted research and interventions to address their specific health concerns (44).

### Impacts of head portering on female head porters

The occupational hazards of head portering among head porters are complex, impacting physical, mental, and social well-being. Due to the strenuous nature of head portering combined with poor living conditions and limited access to healthcare and social protection, female head porters are exposed to a range of physical, mental, and social health risks (40, 41, 43). Heavy load-carrying activities can lead to various health issues, particularly for women and girls with nutritional deficits (29, 46, 54). Head porters often carry loads twice their body weight multiple times a day (37). The metabolic cost of carrying external loads heavier than 15% of one’s body weight contributes to exertion in the legs and chest, increasing oxygen uptake (49), whilst carrying heavy loads without adequate rest and nutrition results in musculoskeletal disorders, injuries, and chronic fatigue (42, 47). Poor living and working conditions, such as long work hours and lack of protective gear, further increase health risks (38). Mentally, stressors like financial instability and discrimination contribute to anxiety and depression within this group (31, 32). Socioeconomic inequalities and limited access to healthcare also heighten the vulnerability of female head porters (4, 9, 19).

In a study of the health impacts of pedestrian head loading in Ghana, Malawi, and South Africa (46), it was reported that head porters experienced headaches, neck and//or back pain, exhaustion, risk for acute injury to the legs, chronic musculoskeletal symptoms, and reproductive health challenges. Porter (2014) also noted the energy cost of head loading on cognitive function and impaired immune function. Injuries are attributed to the size and frequency of the load carrying as well as lack of habituation to increasing load, carrying heavy loads over long distances and navigating rugged terrain (11, 14, 18, 24–26, 27, 29, 34, 35, 37, 39, 50, 51).

Pelvic prolapse and symptoms of lower abdominal discomfort in women who carry heavy loads for work and as part of domestic activities were also reported (25, 33, 35). Additionally, pregnant head porters often reported muscle fatigue and back injury due to the prolonged postural strains caused by the trunk being displaced in pregnant load carriers (15, 16). The informal nature of working as a head porter makes it accessible to all; however, without proper conditioning and training to carry the load safely, this livelihood strategy makes the work unsafe for many women and girls.

A preliminary search of MEDLINE, the Cochrane Database of Systematic Reviews, and JBI Evidence Synthesis did not identify any current or ongoing systematic or scoping reviews on the topic.

## Materials and Methods

This scoping review aims to systematically explore and map the literature on occupational health hazards and their effects on the health of head porters. Synthesising the available evidence, it seeks to identify the range of health outcomes associated with head portering while examining the social, environmental, and economic determinants.

### Review question

This review will address the following question: What is known about the occupational health hazards of head portering and its effects on head porters’ physical, social and mental health?

### Inclusion and Exclusion Criteria

#### Inclusion

##### Participants

Females of all ages in Sub-Saharan African markets who carry commercial loads on their heads for a living.

##### Concept

Report on load-carrying activities and their impacts on the health of female subjects

**1.** Report occupational health hazards of load-carrying activities and their effects on the health of female subjects. We define health as follows: Health is a state of complete physical, mental and social well-being and not merely the absence of disease or infirmity, as per WHO (2024) (https://www.who.int/about/governance/constitution)
**2.** Activities considered in this review are: Head carrying of any load (food items, construction materials, water, firewood, goods, etc.) for commercial purposes only.

#### Exclusion

- If the sole subjects of the article are male,
- published in a language other than English,
- published before 1994 and not substantial to the population,
- published as commentaries and reviews, and
- were conducted outside of sub-Saharan Africa.

#### Timeframe and languages

Timeframe: Published in the last 31 years (between 1994 and 2025). Languages: English

#### Context

Sources included in this review will include literature from settings from any sub-Saharan African country where women and girls work as head porters in urban commercial markets. Participants will be individuals from all ethnic, racial, and religious backgrounds.

#### Types of sources

Several literature sources will be included in this review. The sources to be included are quantitative studies, qualitative studies, mixed method studies, program evaluations, quality improvement reports, dissertations, theses, peer-reviewed conference papers, and opinion pieces that will be considered for inclusion. Unpublished reports will be incorporated in this review. Grey literature will be limited to national and international governmental and non-governmental websites that report on work with women who carry loads for a living, such as the World Health Organization (WHO) and the United Nations Agencies (UN).

We will include articles from the last 31 years focusing on head porters in sub-Saharan Africa, especially related to women’s and girls’ health. This decision is based on the increase in research dedicated to the health of female head porters during this period, which coincides with a significant rise in the number of girls migrating for informal work (45–47). Therefore, we will include studies and relevant reports published between 1994 and 2025.

## Methods

To understand the impact of head porters on the well-being of female migrants, it was necessary to select a scoping review approach that was not only rigorous in methodology but would also permit the inclusion of diverse types of evidence. A scoping review will allow the mapping of existing literature for the identified phenomenon and its characteristics (8). The six stages of the Arksey and O’Malley (2005) framework will be used to guide and report this scoping review. The study methodology also adheres to the PRISMA guidelines for scoping reviews (53). The Kaya Yei Association, a group of head porters located in Ghana, was consulted as a stakeholder entity representing this population to determine the need and relevance of this study.

The review has been registered with Open Science Framework Registries: https://osf.io/c3q7as.

### Search strategy

#### Stage 1-Identifying research questions

The final question refined from the preliminary search is: “What is known about the physical, social and mental health implications of head portering impacting female head porters?”

#### Stage 2-Identifying relevant studies

The search strategy will aim to locate both published and unpublished studies. This review will use a three-step search strategy. First, a limited MEDLINE (OVID) search was undertaken to identify seed articles. The keywords in the titles and abstracts of seed articles and the index terms used to describe the articles were used to develop a complete search strategy (see Appendix 1). The search strategy, including all identified keywords and index terms, will be adapted for each included database and information source.

The keywords in the search include head porters, its synonyms, female and its synonyms, and Africa. In consultation with the librarian (E.L.), we searched the keywords and synonyms to determine the feasibility of the research questions. The initial search with the librarian used keywords and their synonyms. The search on OVID/Medline began with a broad search using all relevant keywords combined with Boolean operators (AND, OR):

1- “head porter* or head-porter* or head carr* or head-carr* or head-load* or pedestrian load* or head load* or load bear* or load carry* or head carry*”
2- “women or woman or female* or girl” OR “women/ or battered women/ or pregnant women/ or women, working/’
3- “Africa”

Searches 1 AND, 2, AND 3 were combined to identify primary articles.

The search will be refined by adding database-specific subject headings and indexing terms. We will review and refine the search strategy iteratively based on the initial search results and feedback from experts in the field. CINAHL, Scopus, Web of Science, Hinari, Global Index, Medicus and Google Scholar will be used to capture titles that might be missed by adapting the keywords and their synonyms to the specific databases. Conference proceedings, websites, search engines like Google or other online resources, and contact with study authors will be included to broaden the search to capture unpublished studies and grey literature. Forward and backward citation searches of references of relevant studies will be screened for additional studies.

Articles will be included in the review if they meet the following criteria:

- The study population include females of any age engaging in head-carrying activities for a living in sub-Saharan Africa
- Report on load-carrying activities and their impact on the health of female subjects.
- Articles including grey literature will be included if published in English.
- We will include articles from the last 30 years focusing on head porters in sub-Saharan Africa, especially related to women’s and girls’ health. This decision is based on the increase in research dedicated to the health of female head porters during this period, which coincides with a significant rise in the number of girls migrating for informal work (Porter, 2014; Porter et al., 2012; 2013). Therefore, we will include studies and relevant reports published between 1994 and 2024.
- The sources included will be quantitative, qualitative, mixed methods, program evaluation articles, quality improvement reports, dissertations/theses, peer-reviewed conference papers, opinion pieces, and unpublished reports, which will be incorporated into this review. The unpublished reports will be limited to national and international, governmental, and non-governmental websites that work with women who carry loads for a living, such as the WHO and the UN.

Articles will be excluded if they meet the following criteria:

- Study subjects are solely male
- Published in a language other than English
- Published before 1994
- Published as commentaries and other reviews
- Source context outside of sub-Saharan Africa.

### Study/Source of Evidence Selection

#### Stage 3-Study selection

Following the search, all identified citations will be collated and uploaded into Covidence, and duplicates will be removed. Following a pilot test, titles and abstracts will be screened by two or more independent reviewers for assessment against the inclusion criteria for the review. Potentially relevant sources will be retrieved in full, and their citation details will be imported into Covidence. Two independent reviewers will assess the full text of selected citations in detail against the inclusion criteria. Reasons for excluding sources of evidence in full text that do not meet the inclusion criteria will be recorded and reported in the scoping review. Any reviewer disagreements at each stage of the selection process will be resolved through a third reviewer. The search results and the study inclusion process will be reported in full in the final scoping review and presented in a PRISMA flow diagram (Tricco et al., 2016). During the completion of this scoping review protocol, the search strategy was implemented to determine the feasibility of the scoping review. The preliminary and final search results will be displayed using the PRISMA Extension for Scoping Review (PRISMA-Scr) in this protocol and the final report. Data extraction, analysis, and reporting of results have not commenced and are anticipated to require approximately nine months.

### Data extraction

#### Stage 4-Charting the data

Data will be extracted from papers included in the scoping review by two independent reviewers using a data extraction tool developed by the reviewers (See Appendix I). The data extracted will include specific details: author (s), year of publication, study type, country of origin, aim/purpose, population characteristics/sample size, gaps identified, study recommendations and key findings relevant to the review question/s. To calibrate reviews initially, all reviewers will review and extract the data for the first 10 papers to ensure consistency and clarity between them.

The draft data extraction tool will be modified and revised as necessary while extracting data from each included evidence source. The modifications will be detailed in the scoping review. The third reviewer will resolve any disagreements between the reviewers. If appropriate, paper authors will be contacted three times to request missing or additional data. If there is no response, the deficit papers will be removed.

Critical appraisal of individual sources of evidence will not be completed, as this is generally not required for scoping reviews. A PRISMA chart will present the decision tree regarding the studies selected for inclusion.

### Data analysis and presentation

#### Stage 5-Collating, summarising, and reporting the results

The extracted data will be reported using a narrative summary to supplement data tables. The findings will inform recommendations for future practice, policy, and research.

#### Stage 6-Consulting, Ethics and Dissemination

Findings from this scoping review will be shared with all stakeholders involved. This study’s findings will be disseminated through community engagement activities, such as community meetings, to discuss the findings and implications. Formal knowledge translation and dissemination activities will include publishing findings in African-based high-impact journals to reach relevant audiences. Academic knowledge translation activities will consist of conference presentations and peer-reviewed journal publications. The final recommendations for practice, policy, and research will be shared and distributed through toolkits for healthcare professionals, policy briefs, and information leaflets.

## Supporting information

Search Strategy

Data extraction table

Prisma Chart

## Data Availability

No datasets were generated or analysed during the current study. All relevant data from this study will be made available upon study completion

## Author contributions

Veronica Segbedzie conceptualised this study and completed the original manuscripts. Dr Solina Richter contributed to the study selection as the second reviewer and completed the revisions and edits to the manuscript. Dr. Pammla Petrucka served as the third reviewer and completed the final revisions and edits to the manuscript.

## Acknowledgements

This scoping review protocol will contribute towards the Doctor of Philosophy in Nursing degree requirements for Veronica Segbedzie PhD(c).

## Metadata

### Funding

The authors did not receive any funding to conduct the scoping review protocol.

### Declarations

The work was completed by female authors who hail from Africa and/or have lived on the continent and, therefore, write about issues directly related to vulnerable and marginalised women populations in Africa.

### Data Availability

This is a scoping review protocol. Currently, no data is available. Research data will be made publicly available when the study is completed and published.

