## Supplementary material for "Health Implications of Head Portering in Female Head Porters: A Scoping Review Protocol": Search Strategy

### Appendix I: Search strategy **Database:**Ovid MEDLINE(R) <1946 to November Week 3 2024>

| **#** | **Query** | **Results from 26 Nov 2024** |
| --- | --- | --- |
| 1 | (head porter* or Kaya yei or kayayei or kaya yoo or pedestrian load* or head load* or load bear* or load carry* or head carry*).ti,ab,kf. | 4,855 |
| 2 | (women or woman or female* or girl).ti,ab,kf. | 2,272,080 |
| 3 | women/ or battered women/ or pregnant women/ or women, working/ | 38,547 |
| 4 | 2 or 3 | 2,281,764 |
| 5 | 1 and 4 | 290 |
| 6 | (head port* or head-port* or Kaya yei or kayayei or kaya yoo or pedestrian load* or pedestrian-load* or head load* or head-load* or load bear* or load-bear* or load carry* or load-carry* or head carry* or head-carry*).ti,ab,kf. | 5,001 |
| 7 | (women or woman or female* or girl).ti,ab,kf. | 2,272,080 |
| 8 | women/ or battered women/ or pregnant women/ or women, working/ | 38,547 |
| 9 | 7 or 8 | 2,281,764 |
| 10 | [africa.mp](http://africa.mp/). [mp=title, book title, abstract, original title, name of substance word, subject heading word, floating sub-heading word, keyword heading word, organism supplementary concept word, protocol supplementary concept word, rare disease supplementary concept word, unique identifier, synonyms, population supplementary concept word, anatomy supplementary concept word] | 180,479 |
| 11 | exp Africa/ | 345,023 |
| 12 | 10 or 11 | 382,066 |
| 13 | 6 and 9 and 12 | 27 |

### (head porter* or Kaya yei or kayayei or kaya yoo or pedestrian load* or head load* or load bear* or load carry* or head carry*).ti,ab,kf. (women or woman or female* or girl).ti,ab,kf. women/ or battered women/ or pregnant women/ or women, working/ 2 or 3 1 and 4 (head port* or head-port* or Kaya yei or kayayei or kaya yoo or pedestrian load* or pedestrian-load* or head load* or head-load* or load bear* or load-bear* or load carry* or load-carry* or head carry* or head-carry*).ti,ab,kf. (women or woman or female* or girl).ti,ab,kf. women/ or battered women/ or pregnant women/ or women, working/ 7 or 8 [africa.mp](http://africa.mp/). [mp=title, book title, abstract, original title, name of substance word, subject heading word, floating sub-heading word, keyword heading word, organism supplementary concept word, protocol supplementary concept word, rare disease supplementary concept word, unique identifier, synonyms, population supplementary concept word, anatomy supplementary concept word] exp Africa/ 10 or 11 6 and 9 and 12
