## Supplementary material for "Health Implications of Head Portering in Female Head Porters: A Scoping Review Protocol": Data extraction table

### Appendix II: Data extraction instrument

| **#** | **Author(s)** | **Title** | **Year of publication** | **Country of origin** | **Aims/purpose** | **Pop/Sample Size** | **Methodology/methods** | **Gaps** | **Recommendation** |
| --- | --- | --- | --- | --- | --- | --- | --- | --- | --- |
| 1 |  |  |  |  |  |  |  |  |  |
